# Anti-SARS-CoV-2 Antibody Levels Associated with COVID-19 Protection in Outpatients Tested for SARS-CoV-2, US Flu VE Network, October 2021–June 2022

**DOI:** 10.1101/2023.09.21.23295919

**Authors:** Kelsey M. Sumner, Ruchi Yadav, Emma K. Noble, Ryan Sandford, Devyani Joshi, Sara Y. Tartof, Karen J. Wernli, Emily T. Martin, Manjusha Gaglani, Richard K. Zimmerman, H. Keipp Talbot, Carlos G. Grijalva, Jessie R. Chung, Eric Rogier, Melissa M. Coughlin, Brendan Flannery

## Abstract

**Background:** We assessed the association between antibody concentration ≤5 days of symptom onset and COVID-19 illness among patients enrolled in a test-negative study

**Methods:** From October 2021─June 2022, study sites in seven states enrolled and tested respiratory specimens from patients of all ages presenting with acute respiratory illness for SARS-CoV-2 infection using rRT-PCR. In blood specimens, we measured concentration of anti-SARS-CoV-2 antibodies against the ancestral strain spike protein receptor binding domain (RBD) and nucleocapsid (N) antigens in standardized binding antibody units (BAU/mL). Percent reduction in odds of symptomatic COVID-19 by anti-RBD antibody was estimated using logistic regression modeled as (1–adjusted odds ratio of COVID-19)x100, adjusting for COVID-19 vaccination status, age, site, and high-risk exposure.

**Results:** A total of 662 (33%) of 2,018 symptomatic patients tested positive for acute SARS-CoV-2 infection. During the Omicron-predominant period, geometric mean anti-RBD binding antibody concentrations measured 823 BAU/mL (95%CI:690─981) among COVID-19 case-patients versus 1,189 BAU/mL (95%CI:1,050─1,347) among SARS-CoV-2 test-negative patients. In the adjusted logistic regression, increasing levels of anti-RBD antibodies were associated with reduced odds of COVID-19 for both Delta and Omicron infections.

**Conclusion:** Higher anti-RBD antibodies in patients were associated with protection against symptomatic COVID-19 during emergence of SARS-CoV-2 Delta and Omicron variants.

## INTRODUCTION

COVID-19 vaccine trials and immunologic studies have evaluated neutralizing antibodies as potential immune correlates of protection from COVID-19 illness [1, 2]. Immunobridging studies have correlated anti-SARS-CoV-2 binding antibody (bAb) levels against ancestral spike and receptor binding domain (RBD) antigens with virus neutralizing antibody levels [3]. Immune correlates of protection following vaccination are important for immunobridging studies and potential evaluation of new COVID-19 vaccines and formulations [1, 2, 4]. Assessing protective antibody levels in the population may help not only vaccine evaluation but also prediction of susceptibility to and protection against emerging variants [5]. Immune correlates are continually re-evaluated as levels of protection mediated by antibodies vary with time and emergence of new SARS-CoV-2 variants.

Observational studies of licensed vaccines can contribute to understanding immune biomarkers associated with protection against COVID-19 illness. Observational, test-negative design (TND) studies are widely used to evaluate influenza and COVID-19 vaccine effectiveness [6, 7] and may be used to estimate antibody levels proximal to illness onset, which may correlate with protection [8]. TND COVID-19 vaccine effectiveness (VE) studies systematically enroll and test symptomatic patients who sought medical care for an acute respiratory illness [7, 9]. Reduction in the odds of laboratory-confirmed illness indicates VE against disease endpoints. Collection of sera or blood spots at enrollment near the onset of illness can be used in serologic assays to measure bAb levels early in infection that may estimate antibody titers close to the time of infection. Anti-RBD are elicited by U.S.-licensed COVID-19 mRNA vaccines but the vaccine does not elicit antibodies against the nucleocapsid (N) protein [10, 11]; thus, the presence of anti-N antibodies can be an indicator of past SARS- CoV-2 infection among vaccinated and unvaccinated individuals and anti-RBD antibodies can be an indicator of infection or vaccine-induced protection. To assess associations between symptomatic COVID-19 and anti-SARS-CoV-2 RBD and N protein immunoglobulin G (IgG) antibody levels, we quantified bAb levels during acute respiratory illness in patients enrolled in a COVID-19 VE study.

## MATERIALS AND METHODS

### Study population and sample collection

Ambulatory patients aged 1 year or older presenting within 10 days of onset of respiratory illness were enrolled from participating healthcare facilities across seven study sites in the US Influenza Vaccine Effectiveness Network, as previously described [12, 13].

Epidemiologic data collected from enrolled patients included patient age, date of illness onset, reported symptoms, documented COVID-19 vaccination history including dates of COVID-19 vaccination, and dates of prior positive COVID-19 tests recorded in electronic medical records. Respiratory specimens (nasal/nasopharyngeal and throat swabs) were tested for SARS-CoV-2 by real-time reverse-transcription polymerase chain reaction (rRT-PCR). Patients were classified based on test results as COVID-19 cases or SARS-CoV-2 test-negative controls.

SARS-CoV-2 variant infection was determined by genomic sequencing or categorized by predominant variant during two time periods as previously described [12–14]: Delta (October 1– December 24, 2021) or Omicron BA.1–5 (December 25, 2021–June 29, 2022).

At enrollment, research staff at each study site collected blood specimens from participants by finger stick and absorbed drops on Whatman 903 filter paper cards. Filter paper blood spots were dried at room temperature, packed with desiccant, and sent to the US Centers for Disease Control and Prevention (CDC). An acute blood specimen had to be collected from a patient within 5 days of symptom onset for inclusion in the analysis (**Supplemental figure 1**).

This activity was reviewed and approved by CDC and each US Flu VE Network site’s Institutional Review Board.

### Serologic assays

Dried blood spots (DBS) have been shown to provide similar results to venipuncture for SARS-CoV-2 antibody testing [15–17]. The FlexImmArray^TM^ SARS-CoV-2 Human IgG Antibody test (Tetracore, Rockville, MD) was used to detect SARS-CoV-2 antibodies in DBS; this test uses a 7-Plex microsphere based assay designed for specific IgG antibody detection against SARS-CoV-2 [18]. It employs three immobilized SARS-CoV-2 recombinant antigens and includes three external sample controls for assessing performance. Diluted samples (1:300) were incubated with the 7-Plex microsphere mixture, and fluorescent anti-human IgG-phycoerythrin was used as the reporter. Readings were obtained using the Luminex MAGPIX instrument (Luminex Corporation, Austin, TX), and result interpretation relied on the manufacturer-provided seropositivity threshold values, represented as median fluorescence intensity (MFI) ratios ≥1.2. To normalize raw MFI values, each test specimen’s MFI value was divided by the mean calibrator MFI value. MFI ratios were standardized and calibrated against the World Health Organization (WHO) anti-SARS-CoV-2 immunoglobulin binding antibody unit (BAU) international standard (20/150) using linear regression [19]. Blinded panels of 30 DBS specimens, including SARS-CoV-2 positive and negative samples, underwent rigorous in-house verification at two sites, confirming the suitability of DBS for SARS CoV-2 human IgG detection with the Tetracore FlexImmArray kit. Antibody concentration in BAU/mL was multiplied by a dilution factor of 300 for analyses (anti-RBD seropositivity cutoff 15.9 BAU/mL; anti-N seropositivity cutoff 6.9 BAU/mL).

### Statistical analysis

Analyses were restricted to patients with a known date of specimen collection and SARS-CoV-2 rRT-PCR result. Patients who received only one dose or >4 doses of COVID-19 mRNA vaccine, any non-mRNA vaccine dose, or a dose of unknown COVID-19 vaccine type were excluded. Demographic characteristics, COVID-19 vaccination status, and prior SARS-CoV-2 infection history were compared between patients testing SARS-CoV-2-positive at enrollment versus patients who tested negative. Geometric mean anti-RBD and anti-N antibody concentration (GMC) was compared across patients by current infection status, COVID-19 vaccination status (unvaccinated, two, three, or four doses), and prior SARS-CoV-2 infection status, defined as electronic medical record documentation of one or more prior positive SARS-CoV-2 tests or anti-N bAb levels in acute sera indicative of prior infection (≥6.9 BAU/mL). Prior infection was documented from March 17, 2020, to June 12, 2022. Distributions of anti-RBD and anti-N bAb levels were plotted by COVID-19 case and test-negative control status and number of COVID-19 vaccines received.

Odds of acute COVID-19 positive cases versus test-negative controls were estimated by anti-RBD antibody level (modeled linearly based on a functional form assessment) using a logistic regression model adjusted for COVID-19 vaccination status (modeled categorically as two, three, or four doses versus unvaccinated), age (modeled with cubic terms based on a functional form assessment), study site, illness onset week, and high-risk SARS-CoV-2 exposure (healthcare worker or contact of lab-confirmed COVID-19 case). Model covariates were defined *a-priori* as previously described [12, 13] and tested for inclusion if they created >5% change in the main effect estimate or significantly improved model fit by the log-likelihood ratio test (**Supplemental Table 1**). Percent reduction in the odds of symptomatic COVID-19 was calculated as (1-adjusted odds ratio) x 100. Models were run stratified by COVID-19 variant period as well as COVID-19 vaccination status.

Next, we evaluated the likelihood of COVID-19 illness by anti-N bAb levels, COVID-19 vaccination status, and evidence of prior SARS-CoV-2 infection. To do so, we compared the number of COVID-19 cases that occurred stratified by anti-N antibody concentration (low <10 BAU/mL, medium 10–99 BAU/mL, and high ≥100 BAU/mL), where higher anti-N antibody concentration reflected more recent prior SARS-CoV-2 infection. These cut points were chosen based on natural inflection points in the functional form of anti-N antibody concentration. The percentage of participants who had anti-RBD bAb concentrations above the threshold for a 50% reduction in odds of symptomatic COVID-19 was estimated stratified by anti-N bAb categorizations, COVID-19 vaccine doses, and evidence of prior SARS-CoV-2 infection.

Anti-RBD and anti-N bAb concentrations were also compared in paired acute and convalescent specimens for a subset of COVID-19 cases. Paired analyses were restricted to SARS-CoV-2 positive patients with known acute and convalescent blood specimen collections within 21–56 days of one another. Anti-RBD and anti-N bAb GMC was calculated in acute and convalescent specimens. The percentage of participants who had anti-RBD bAb concentrations above the 50% threshold for reduction in symptomatic COVID-19 was estimated in the acute and convalescent specimens. All statistical analyses were performed using R version 4.0.3 (R Foundation for Statistical Computing, Vienna, Austria).

## RESULTS

A total of 2,018 enrollees in the US Flu VE network had blood specimens collected at enrollment and were included in analyses (**Figure 1**); 662 (33%) enrollees were COVID-19 case patients and 1,356 (67%) were test-negative patients (**Figure 1**). SARS-CoV-2 positivity varied by variant period; 87 (17%) of 503 patients enrolled during the Delta variant-predominant period tested positive for SARS-CoV-2 and 575 (38%) of 1,515 tested positive during the Omicron-predominant period (**Figure 1, Table 1**). A lower percentage of cases had evidence of SARS-CoV-2 infection prior to their current acute illness in the Delta-predominant period (20%) compared to the Omicron-predominant period (42%) (**Table 1**).

**Figure 1.**
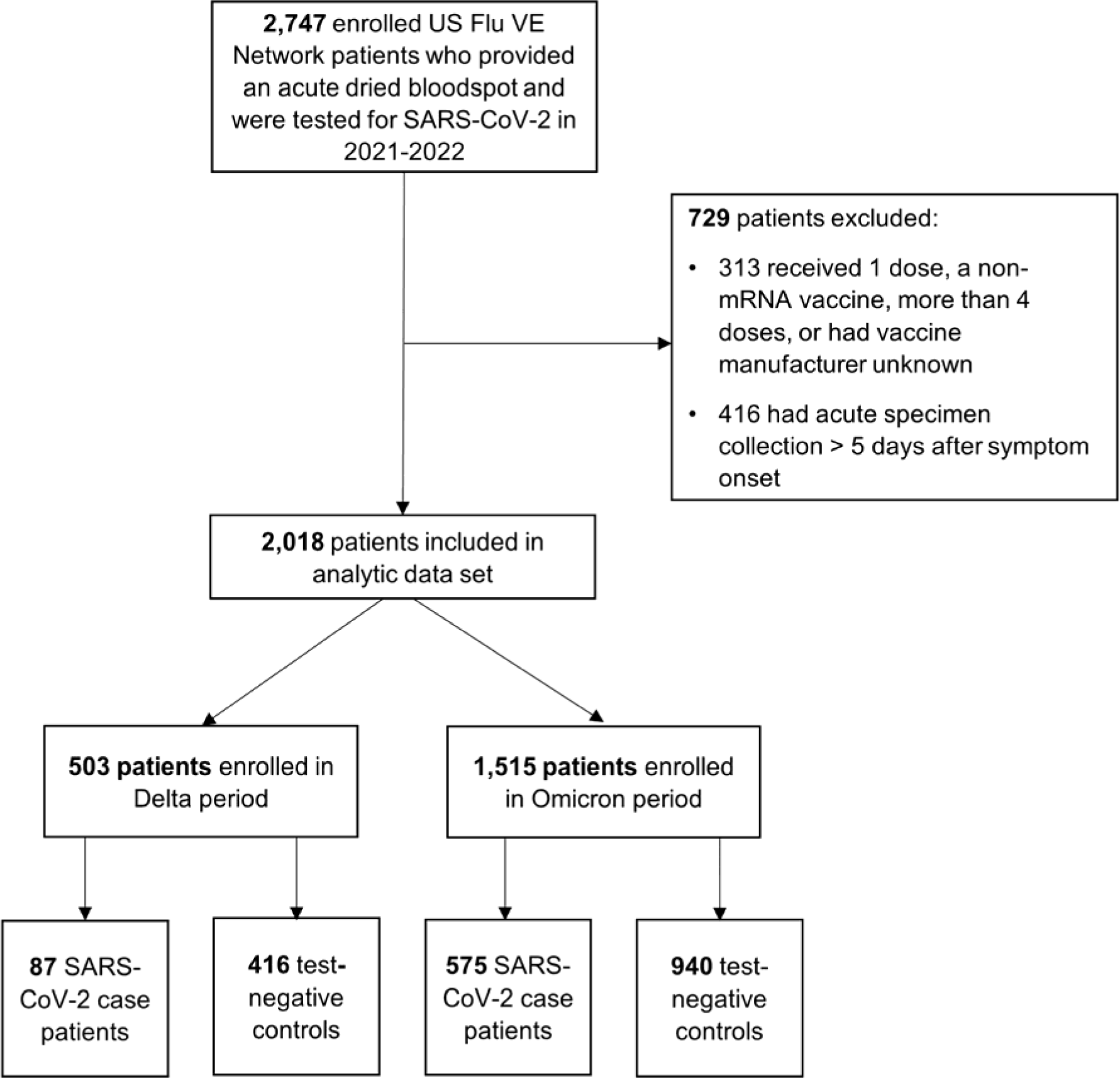
US Influenza Vaccine Effectiveness Network enrollment for 2021–22 season. The number of patients enrolled in the US Flu VE Network and included in the final analytic data set are shown, detailing each of the exclusion criterion applied. The Delta-predominant period was from October 1–December 24, 2021, and the Omicron-predominant period from December 25, 2021–June 29, 2022.

**Table 1.** Characteristics of COVID-19 case and SARS-CoV-2 test negative control patients with acute respiratory illness by SARS-CoV-2 molecular test result.

| Characteristic | Delta period<sup>1</sup> |  |  | Omicron period<sup>2</sup> |  |  |
| --- | --- | --- | --- | --- | --- | --- |
|  | Test-negative control<br>N = 416 | Case<br>N = 87 | p-value<sup>3</sup> | Test-negative control<br>N = 940 | Case<br>N = 575 | p-value<sup>4</sup> |
| Age, Median (range) | 34 (1, 83) | 38 (7, 72) | 0.699 | 38 (3, 82) | 40 (5, 82) | 0.006 |
| Sex<sup>5</sup>, n (%) |  |  | 0.779 |  |  | 0.245 |
| Female | 250 (60%) | 51 (59%) |  | 623 (66%) | 364 (63%) |  |
| Male | 165 (40%) | 36 (41%) |  | 316 (34%) | 210 (37%) |  |
| Race/ethnicity<sup>6</sup>, n (%) |  |  | 0.240 |  |  | 0.009 |
| White, non-Hispanic | 250 (60%) | 54 (64%) |  | 597 (64%) | 321 (57%) |  |
| Black, non-Hispanic | 18 (4.3%) | 6 (7.1%) |  | 38 (4.1%) | 23 (4.1%) |  |
| Asian, non-Hispanic | 38 (9.2%) | 7 (8.2%) |  | 69 (7.4%) | 71 (13%) |  |
| Other, non-Hispanic | 16 (3.9%) | 6 (7.1%) |  | 37 (4.0%) | 26 (4.6%) |  |
| Hispanic | 92 (22%) | 12 (14%) |  | 191 (20%) | 123 (22%) |  |
| Days from symptom onset, Median (range) | 3 (0, 5) | 2 (0, 5) | 0.239 | 2 (0, 5) | 2 (0, 5) | <0.001 |
| COVID-19 vaccination status, n (%) |  |  | 0.029 |  |  | 0.660 |
| Unvaccinated | 82 (20%) | 28 (32%) |  | 160 (17%) | 85 (15%) |  |
| Two doses | 234 (56%) | 45 (52%) |  | 233 (25%) | 150 (26%) |  |
| Three doses | 100 (24%) | 14 (16%) |  | 521 (55%) | 326 (57%) |  |
| Four doses | 0 (0%) | 0 (0%) |  | 26 (2.8%) | 14 (2.4%) |  |
| Evidence of prior SARS-CoV-2 infection<sup>7</sup>, n (%) |  |  | 0.002 |  |  | <0.001 |
| No | 262 (63%) | 70 (80%) |  | 345 (37%) | 333 (58%) |  |
| Yes | 154 (37%) | 17 (20%) |  | 595 (63%) | 242 (42%) |  |
| Self-reported presence of ≥1 medical condition<sup>8</sup>, n (%) | 92 (23%) | 23 (28%) | 0.335 | 257 (28%) | 142 (25%) | 0.243 |
| Healthcare worker or close contact with confirmed COVID-19 case, n (%) | 115 (28%) | 41 (47%) | <0.001 | 406 (43%) | 314 (55%) | <0.001 |
| Study site, n (%) |  |  | 0.046 |  |  | <0.001 |
| California | 194 (47%) | 37 (43%) |  | 243 (26%) | 207 (36%) |  |
| Pennsylvania | 93 (22%) | 29 (33%) |  | 132 (14%) | 57 (9.9%) |  |
| Tennessee | 52 (12%) | 8 (9.2%) |  | 83 (8.8%) | 46 (8.0%) |  |
| Texas | 50 (12%) | 4 (4.6%) |  | 201 (21%) | 83 (14%) |  |
| Wisconsin | 27 (6.5%) | 9 (10%) |  | 72 (7.7%) | 27 (4.7%) |  |
| Michigan | 0 (0%) | 0 (0%) |  | 76 (8.1%) | 51 (8.9%) |  |
| Washington | 0 (0%) | 0 (0%) |  | 133 (14%) | 104 (18%) |  |

<sup>1</sup> The Delta-predominant period was defined as the period from October 1–December 24, 2021.
<sup>2</sup> The Omicron-predominant period was defined as the period from December 25, 2021–June 29, 2022.
<sup>3</sup> Wilcoxon rank sum test; Pearson's Chi-squared test; Fisher's exact test
<sup>4</sup> Wilcoxon rank sum test; Pearson's Chi-squared test
<sup>5</sup> 3 missing sex
<sup>6</sup> 23 missing race/ethnicity
<sup>7</sup> Evidence of prior SARS-CoV-2 infection was defined as electronic medical record documentation of prior positive SARS-CoV-2 tests or anti-N bAb levels in acute sera indicative of prior infection (≥6.9 BAU/mL). Prior infection was documented from March 17, 2020, to June 12, 2022.
<sup>8</sup> 63 missing self-reported presence of ≥1 medical condition

Similar to the Delta-predominant period, during the Omicron-predominant period, GMCs of anti-RBD and anti-N bAb were lower among COVID-19 cases (anti-RBD 822.7 95% CI: 689.9─981.1; anti-N 5.7 BAU/mL 95% CI: 5.0─6.5) compared to test-negative controls (anti-RBD 1189.0 95% CI: 1049.7─1346.8; anti-N 15.5 BAU/mL 95% CI: 13.6─17.8) (**Table 2**). Anti-RBD GMC was higher with increasing number of COVID-19 vaccine doses but decreased with increasing time since COVID-19 vaccination (**Table 2, Supplemental Figure 2**). During both variant periods, anti-RBD GMC was higher in patients with evidence of prior SARS-CoV-2 infection compared to those without evidence of prior infection (**Table 2**).

**Table 2.**
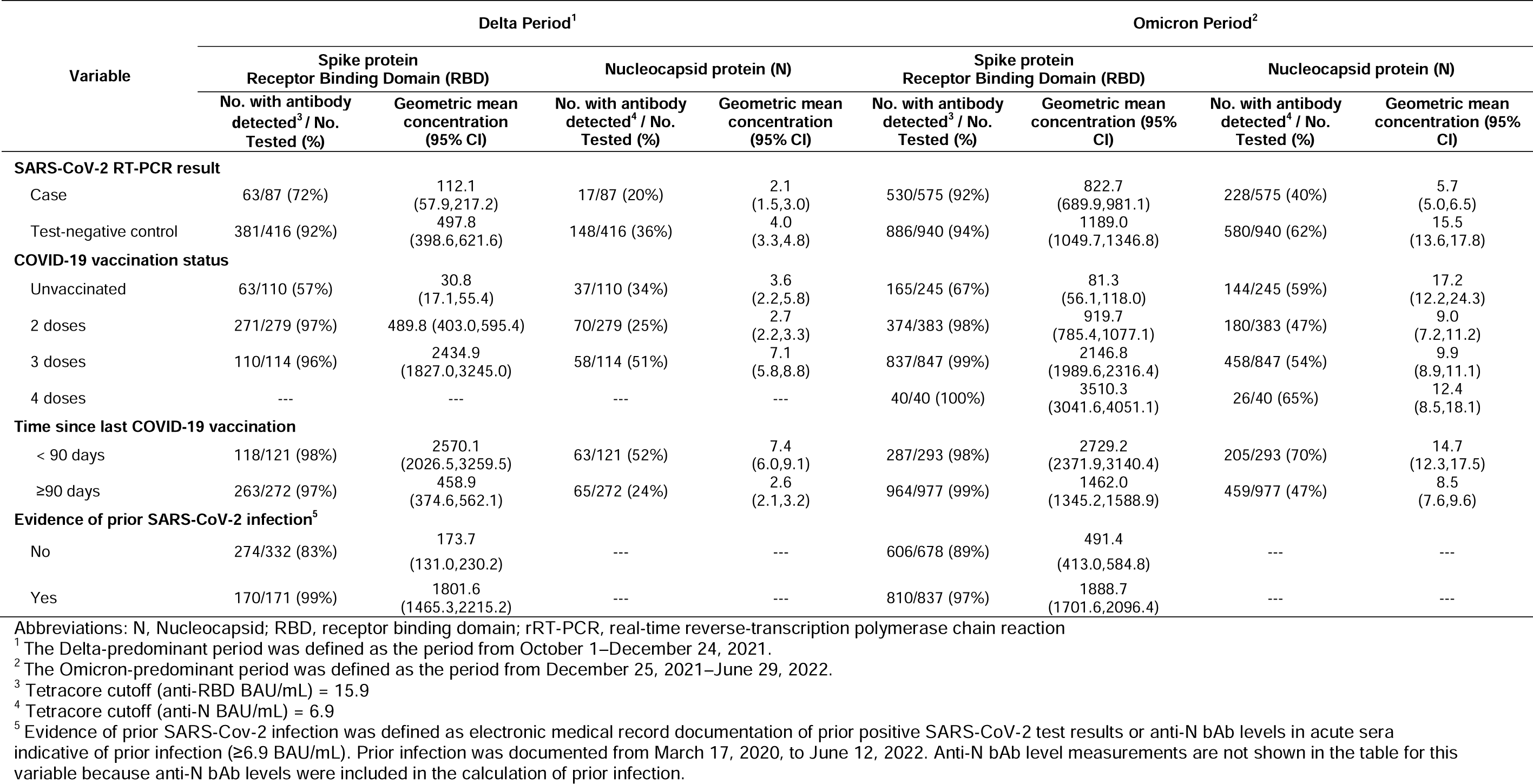
Geometric mean concentrations (BAU/mL) in acute phase dried blood spots for anti-SARS-CoV-2 RBD and N antigens by COVID-19 case status, COVID-19 vaccination status and prior laboratory-confirmed SARS-CoV-2 infection.

During both variant periods, the likelihood of being a symptomatic COVID-19 case decreased with increasing RBD bAb level (**Figure 2A**). Threshold anti-RBD bAb levels associated with 50% reduction in odds of symptomatic COVID-19 were lower during the Delta variant-predominant period (1,968 BAU/mL) than during the Omicron-predominant period (3,375 BAU/mL; **Figure 2B**). There was no clear trend in anti-RBD bAb levels associated with a 50% reduction in odds of COVID-19 when results were stratified by doses of COVID-19 vaccine received (**Supplemental Table 2**). Regardless of variant period, a higher percentage of participants with moderate anti-N bAb levels had anti-RBD bAb levels above the 50% threshold compared to participants with high anti-N bAb levels (**Table 3**). The percentage of participants above the 50% anti-RBD bAb threshold rose with increasing numbers of COVID-19 vaccine doses and evidence of prior SARS-CoV-2 infection (**Table 3**).

**Figure 2.**
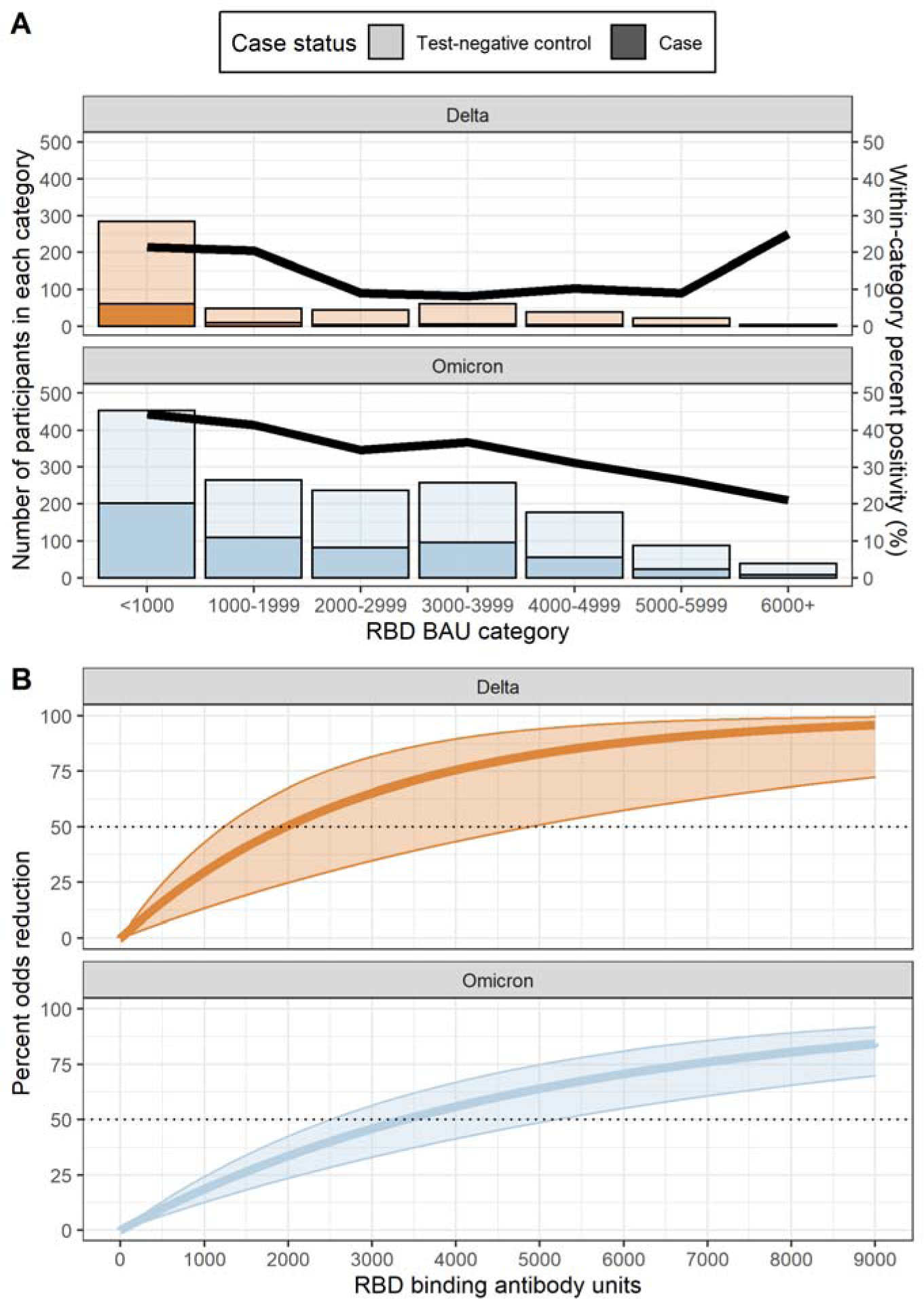
Association between SARS-CoV-2 anti-spike receptor binding domain (RBD) IgG antibodies and likelihood of symptomatic COVID-19. 2A. Bars indicate the number of COVID-19 case (darker shading) and test-negative control (lighter shading) patients within each anti-RBD binding antibody unit (BAU) category. The line represents SARS-CoV-2 rRT-PCR test positivity within each anti-RBD binding antibody category. Results presented stratified by the Delta (orange) and Omicron (grey) variant periods. The Delta-predominant period was from October 1–December 24, 2021, and the Omicron-predominant period from December 25, 2021– June 29, 2022. **2B.** The percent odds reduction in COVID-19 illness by anti-RBD binding antibody level is presented stratified by the Delta (orange) and Omicron (grey) variant periods. Percent odds reduction was estimated as (1-adjusted odds ratio) x 100, using the adjusted odds ratio produced by a logistic regression model adjusted for COVID-19 vaccination status, age, study site, illness onset week, and high-risk SARS-CoV-2 exposure.

**Table 3.**
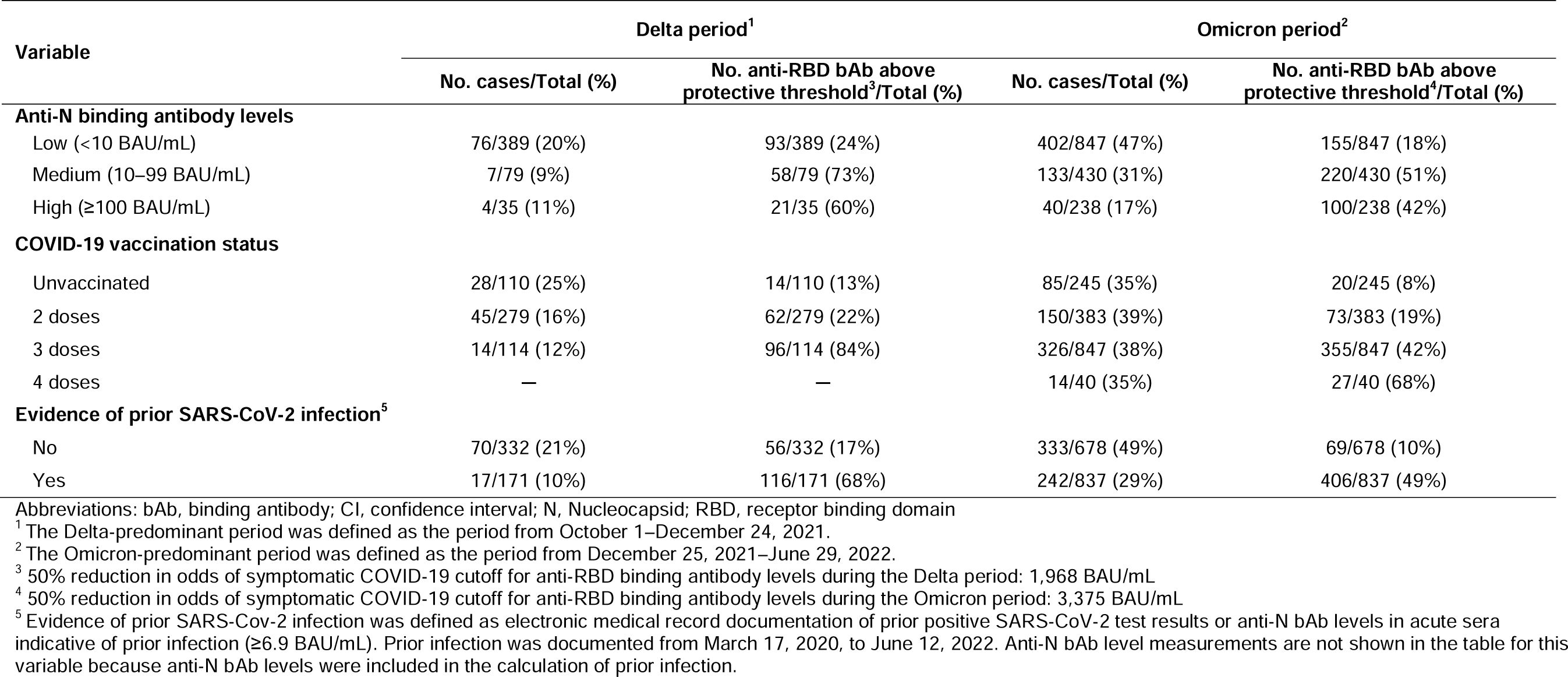
Likelihood of symptomatic COVID-19 by anti-N binding antibody levels, COVID-19 vaccination status, and prior SARS-CoV-2 infection history.

A total of 105 patients with acute SARS-CoV-2 infection and blood specimens collected at both enrollment and convalescent visits were included in an analysis subset; 104 patients were enrolled during the Omicron period and 1 during the Delta period. For the 104 patients enrolled during the Omicron variant period, the GMCs for anti-RBD bAb in acute and convalescent sera were 1257.8 (95% CI: 923.9–1712.3) and 3188.5 (95% CI: 2638.7–3853.0), and the GMCs for anti-N bAb in acute and convalescent sera were 5.5 (95% CI: 4.3–7.1) and 259.4 (95% CI: 200.6–335.4), respectively; at the acute phase, 27 (26%) patients had anti-RBD bAb levels above the threshold that correlated to a 50% reduction in odds of acute SARS-CoV-2 infection (3,375 BAU/mL) compared to 73 (72%) patients during the convalescent phase that had anti-RBD bAb levels above this threshold (results not shown).

## DISCUSSION

In this observational study of patients with acute respiratory illness, the likelihood of symptomatic COVID-19 decreased as levels of bAb against ancestral SARS-CoV-2 spike RBD antigen increased. During acute-phase illness, anti-RBD bAb concentrations of 1,968 BAU/mL corresponded to a 50% reduction in the likelihood of COVID-19 illness during the Delta-predominant period compared to low anti-RBD bAb levels. During the Omicron-predominant period, a 50% reduction in COVID-19 illness was observed at 3,375 BAU/mL. Patients with the highest anti-RBD antibody levels did not always have the highest anti-N antibody levels.

Using SARS-CoV-2 test-negative control patients, we found that a higher concentration of anti-RBD IgG antibodies was correlated with decreased odds of COVID-19 illness [20].

Studies from COVID-19 vaccine trials have correlated anti-SARS-CoV-2 bAb levels against ancestral spike and RBD antigens with virus neutralizing antibody levels, which likely play a key role in protection [3]. The test-negative design provides efficient enrollment of patients with laboratory-confirmed illness (depending on proportion of COVID-19 illness among patients seeking care) and an uninfected comparison group of patients seeking care for similar illness [8, 21]. While distributions of anti-RBD IgG antibody concentrations in the current study largely overlapped between COVID-19 cases and test-negative controls, higher antibody levels were associated with a reduced likelihood of COVID-19 illness. We observed that over 70% of case patients reached an anti-RBD bAb level above the threshold to reduce the odds of infection by 50% in their convalescent sera. These results suggest that test-negative studies may provide a means of estimating correlates of protection as new SARS-CoV-2 variants emerge.

This study demonstrated that patients with high anti-RBD bAb levels did not always have high anti-N levels. This observed difference could be due to differences in prior vaccination and infection history, as U.S.-licensed COVID-19 mRNA vaccines elicit anti-RBD but not anti-N antibodies [10, 11]; anti-N antibody responses to infection have been observed to differ among vaccinated versus unvaccinated persons [22] and non-neutralizing antibody mechanisms of protection mediated by anti-N bAb may be less affected by variants. We were unable to assess time from most recent SARS-CoV-2 infection for all patients; however, anti-N antibody levels may also reflect a shorter time interval between prior and current SARS-CoV-2 infection. Future analyses could further assess the role anti-N bAbs play in protection against SARS-CoV-2 infection.

The current analysis was aided by the collection of DBS from symptomatic patients at the time of clinical presentation. In a previous analysis, presence of anti-N antibody in acute phase blood spot specimens classified five times as many patients with prior SARS-CoV-2 infection as self-reported or electronic medical record documented COVID-19 [12, 13]. DBS were recognized early in the COVID-19 pandemic as alternatives to venous blood collection for anti-SARS-CoV-2 binding assays [16, 23, 24]. Self-collected DBS that could be shipped by mail facilitated SARS-CoV-2 seroprevalence [25–32] and longitudinal household cohort studies [27, 33]. The binding antibody concentrations against ancestral RBD and N antigens observed using acute-phase DBS were consistent with those measured at illness onset and the convalescent phase using the Meso Scale Diagnostics quantitative binding assay utilized in many COVID-19 vaccine trials [1, 3, 5, 21]. Studies evaluating bAb from DBS and serum specimen types may provide additional tools for evaluating correlates of protection against future SARS-CoV-2 variants [2, 5, 34].

These findings are subject to several limitations. First, results are limited to mild-to-moderate ambulatory illness. Immune markers associated with protection against severe disease should be investigated. In addition, except for the subset of 105 paired acute and convalescent samples, DBS used in this study were collected at one time point during acute illness. Acute-phase antibody titers may reflect early antibody rise in some individuals, resulting in an overestimation of the antibody response at the time of infection for cases; however, analyses were restricted to acute-phase specimens collected within 5 days of symptom onset to limit influence of early antibody response as much as possible. Antibody levels were assessed against ancestral RBD and N antigens rather than against antigens representative of SARS-CoV-2 variants circulating at the time of infection. Levels of variant-specific antibody associated with protection are likely lower than those measured using ancestral antigens. A low number of individuals received four COVID-19 vaccine doses, and small sample sizes limited our ability to compare 50% thresholds for a reduction in odds of COVID-19 stratified by vaccine doses, with results inconsistent. Further, use of DBS in this multi-plex, microsphere assay was previously validated against qualitative serologic assays [18] but not against standardized assays widely used to quantify bAb levels [35]. This study was designed to assess applicability of the test-negative study design to interrogate antibody levels associated with SARS-CoV-2 associated illness. Validation of specimen types and serologic assays is needed before acceptance of this approach.

Overall, these results suggest a role for observational studies designed to assess vaccine effectiveness in evaluating immune correlates of protection. Standardization of serologic assays and ongoing immunobridging studies using well-characterized sera will be needed to update correlates of protection against circulating SARS-CoV-2 variants and facilitate approval of new vaccines [4]. With multiple licensed and recommended COVID-19 vaccines, observational studies incorporating immune markers can complement immunogenicity studies in evaluation of relative vaccine effectiveness.

## ACKNOWLEDGMENTS

We acknowledge Edward A. Belongia from the Marshfield Clinic; Bruno Lewin, Ana Florea, Jennifer Ku, Vennis Hong, Harp Takhar, Sally Shaw, Jeniffer Kim, Britta Amundsen, Ashley McDaniel, Raul Calderon, Gabriela Jimenez, Alicia Torres, Alexandria Reyes, Korina Chen, and Susie Flores from Kaiser Permanente Department of Health Research Science; C. Hallie Phillips, Erika Kiniry, Stacie Wellwood, Kathryn Moser, Brianna Wickersham, Matt Nguyen, Rachael Doud, Suzie Park from Kaiser Permanente Washington Health Research Institute; Dayna Wyatt, Stephanie Longmire, Meredith Denny, Zhouwen Liu, and Yuwei Zhu of Vanderbilt University Medical Center; and Sara S. Kim and Manish Patel of US CDC.

## DISCLAIMER

The findings and conclusions in this report are those of the authors and do not necessarily represent the views of the U.S. Centers for Disease Control and Prevention (CDC). Some authors are federal employees of the United States government, and this work was prepared as part of their official duties. Title 17 U.S.C. 105 provides that ‘copyright protection under this title is not available for any work of the United States Government.’

## DATA AVAILABILITY

All data produced in the present study are available upon reasonable request to the corresponding author.

## SUPPLEMENTAL MATERIALS

**Supplemental Figure 1.**
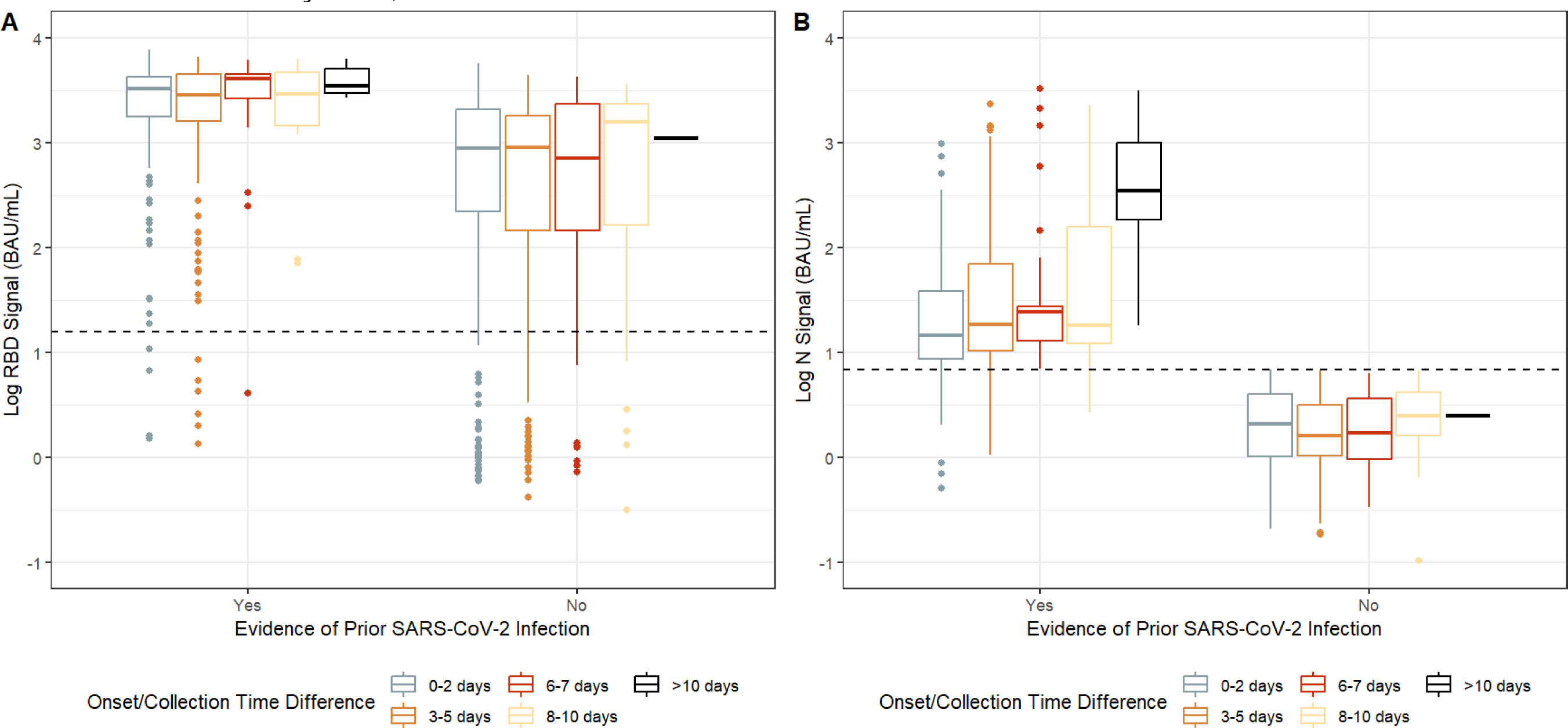
D**i**stribution **of anti-RBD and anti-N binding antibody levels across time between symptom onset and dried blood spot collection.** Anti-RBD (**S1A**) and anti-N (**S1B**) antibody levels (BAU/mL) by days after reported symptom onset among SARS-CoV-2 rRT-PCR positive patients with and without evidence of prior SARS-CoV-2 infection. Binding antibody levels are presented on the log_10_ scale. The dotted line represents the manufacturer’s cutoff for positivity (≥15.9 BAU/mL for anti-RBD and ≥6.9 BAU/mL for anti-N antibody levels).

**Supplemental Figure 2.**
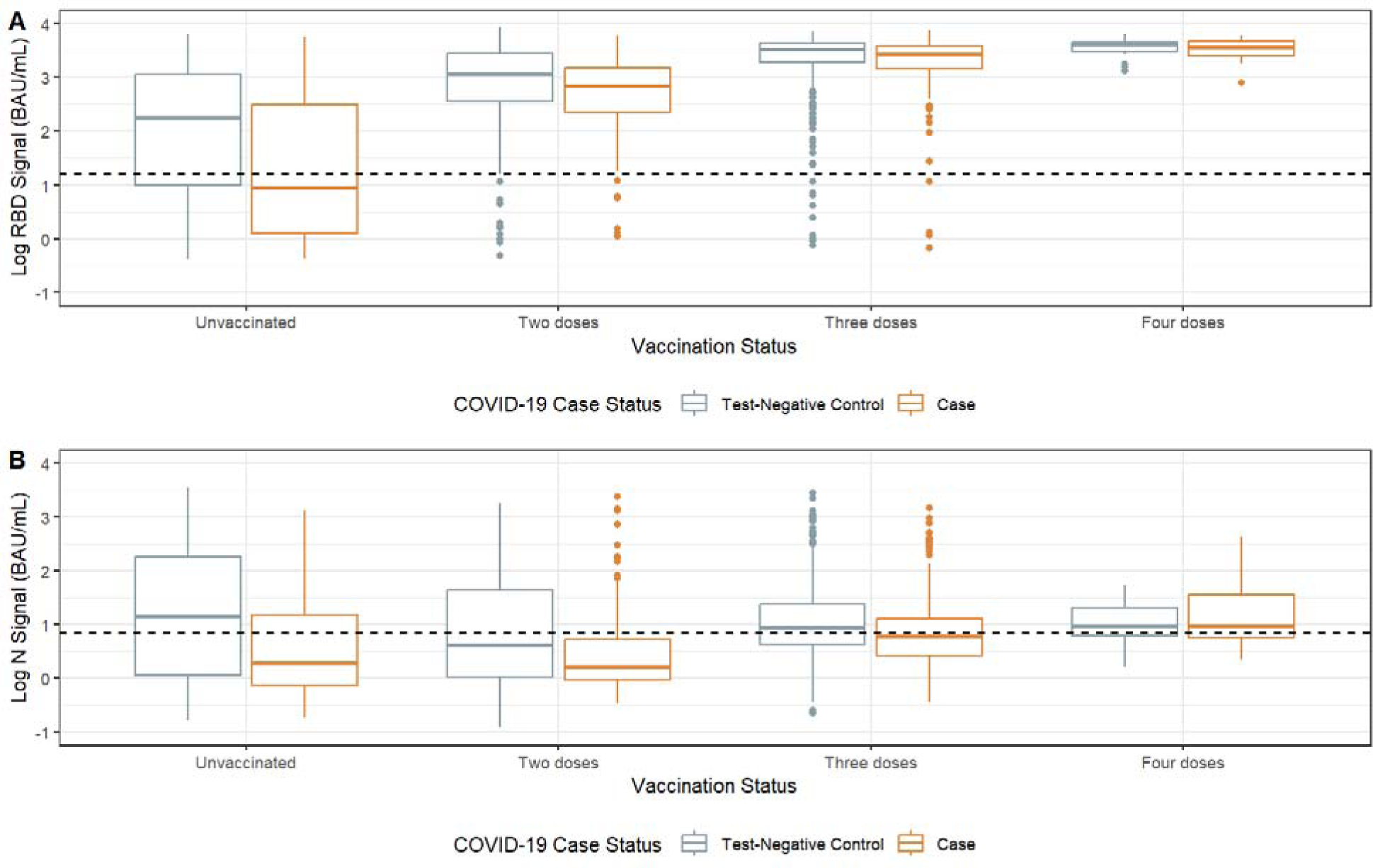
D**i**stribution **of anti-RBD and anti-N binding IgG antibody levels across COVID-19 case and vaccination status.** Anti-RBD (**S2A**) and anti-N (**S2B**) antibody levels (BAU/mL) COVID-19 case and test-negative control status and number of COVID-19 vaccine doses received. Binding antibody levels are presented on the log_10_ scale. The dotted line represents the manufacturer’s cutoff for positivity (≥15.9 for anti-RBD and ≥6.9 for anti-N antibody levels).

**Supplemental Table 1.** Assessment of potential confounding covariates for association between anti-RBD binding antibody levels and symptomatic COVID-19 during the Omicron variant period.

| Covariate assessed | Total number of observations | $\beta^1$ | Percent change in estimate from original adjustment set<sup>2</sup> | AIC | Log-likelihood test $p$-value<sup>2</sup> |
| --- | --- | --- | --- | --- | --- |
| Original adjustment set<sup>3</sup> | 1448 | $-2.1 \times 10^{-4}$ | — | 1826.7 | — |
| Variables removed from original adjustment set: |  |  |  |  |  |
| - **COVID-19 vaccination status** | 1448 | $-2.3 \times 10^{-4}$ | **9.0%** | 1824.1 | 0.340 |
| - **Age (cubed)** | 1448 | $-2.2 \times 10^{-4}$ | 4.3% | 1845.9 | **<0.001** |
| - Sex | 1450 | $-2.1 \times 10^{-4}$ | 0.1% | 1826.7 | NA<sup>4</sup> |
| - Race-ethnicity | 1466 | $-2.1 \times 10^{-4}$ | 0.9% | 1851.3 | NA<sup>4</sup> |
| - **Study site** | 1448 | $-2.0 \times 10^{-4}$ | 3.9% | 1848.6 | **<0.001** |
| - **Illness onset week** | 1448 | $-2.0 \times 10^{-4}$ | **5.4%** | 1836.5 | **<0.001** |
| - Presence of chronic medical condition | 1494 | $-2.0 \times 10^{-4}$ | 4.5% | 1891.5 | NA<sup>4</sup> |
| - **High-risk SARS-CoV-2 exposure** | 1448 | $-2.1 \times 10^{-4}$ | 1.0% | 1837.9 | **<0.001** |

Abbreviations: AIC, Akaike information criterion
<sup>1</sup> The $\beta$ estimate for the association between anti-RBD binding antibody levels (coded linearly) and odds of COVID-19 illness is shown.
<sup>2</sup> Covariates that when added changed the $\beta$ estimate by >5% or had a $p$ -value <0.05 by the log-likelihood ratio test are bolded.
<sup>3</sup> Original adjustment set included COVID-19 vaccination status, age, sex, race-ethnicity, study site, illness onset week, presence of at least one chronic medical condition, and high-risk SARS-CoV-2 exposure.
<sup>4</sup> Not applicable because different total number of observations than adjusted model.

**Supplemental Table 2.** Fifty percent threshold for reduced odds of symptomatic COVID-19 stratified by doses of COVID-19 vaccine received.

| Variable | Delta period<sup>1</sup> |  | Omicron period<sup>2</sup> |  |
| --- | --- | --- | --- | --- |
|  | No. cases/Total (%) | 50% threshold (BAU/mL)<sup>3</sup> | No. cases/Total (%) | 50% threshold (BAU/mL)<sup>3</sup> |
| **COVID-19 vaccination status** |  |  |  |  |
| Unvaccinated | 28/110 (25%) | 696 | 85/245 (35%) | 2943 |
| 2 doses | 45/279 (16%) | 3128 | 150/383 (39%) | 1712 |
| 3 doses | 14/114 (12%) | 961 | 326/847 (38%) | 8528 |
| 4 doses | — | — | 14/40 (35%) | 2680 |

Abbreviations: N, Nucleocapsid; RBD, receptor binding domain
<sup>1</sup> The Delta-predominant period was defined as the period from October 1–December 24, 2021.
<sup>2</sup> The Omicron-predominant period was defined as the period from December 25, 2021–June 29, 2022.
<sup>3</sup> Percent reduction in symptomatic COVID-19 was estimated by $(1 - \text{adjusted odds ratio}) \times 100$ , using the adjusted odds ratio produced by a logistic regression model adjusted for age, study site, illness onset week, and high-risk SARS-CoV-2 exposure. The 50% threshold was where the percent reduction curve crossed 50%.

